# *Cold spot*s of postacute rehabilitation therapy delivery among Original Medicare beneficiaries across the United States: Statistical spatial clustering analysis

**DOI:** 10.64898/2026.07.29.26359230

**Authors:** Tiago S Jesus, Mech Frazier, Pedro C Monteiro, Catia S Pinho, Grace K Delaney, Allen W Heinemann, Anne Deutsch

## Abstract

This study aims to map significant *cold spots* of postacute rehabilitation therapy delivery rates for Original Medicare beneficiaries in the U.S. and determine the prevalence of those *cold spots* in rural areas. Statistical spatial clustering of postacute therapy delivery rates was conducted in ArcGIS Pro using *hot* and *cold spot* analyses (Getis-Ord Gi*). County-level therapy delivery volume was defined as the total minutes of physical, occupational, and speech therapy provided by skilled nursing facilities (SNFs), home health agencies (HHAs), and inpatient rehabilitation facilities (IRFs). Therapy delivery rates were then computed as minutes per Original Medicare beneficiary at the county level and adjusted using a county-level Hierarchical Condition Category risk score. Spatial clustering identified *cold spots* (statistically significant clusters of low rates) and hot spots (clusters of high rates). We also computed the proportion of *cold spots* in rural counties and the relative percentage difference compared to the national rural county baseline, using two rural classification systems. Identified *cold spots* varied by provider type. For SNFs, they were notably identified in the Mountain and West North Central US divisions. For HHAs, *cold spots* appeared across more U.S. Census Divisions, including areas (e.g., Kentucky, Indiana, southern Illinois) where SNFs showed *hot spots*. *Cold spots* were more prevalent in *rural*—and especially in *small rural*—counties across all provider types. In *rural* counties, cold spot rates were 42.8% to 71.5% higher than the rural county baseline. In *small rural* counties, differences were larger, at 69.1% to 95.5% higher. Concluding, *cold spots* of postacute therapy delivery varied across the continental U.S. by provider type but were more prevalent in rural and especially in rural counties with smaller population size ─ across provider types. Identifying these *cold-spot* locations may support geographically targeted policy responses and the development of alternative service-delivery models in underserved areas.

## Introduction

Following an acute care hospitalization for a major illness or injury (e.g., after a stroke, hip fracture), postacute rehabilitation services are often required to address disability in older patients, supporting functional improvement and effective transitions to the home or community[1]. Among Medicare beneficiaries in the USA, more than 40% of acute-hospital patients were discharged to a postacute care provider[2]. Within postacute care settings, rehabilitation services can be provided in: a) inpatient rehabilitation facilities (IRFs), which provide intensive rehabilitation therapy; b) skilled nursing facilities (SNFs), which generally provide less-intensive inpatient rehabilitation; and c) home health agencies (HHAs), which provide home-based care that may include rehabilitation therapies[2].

Geographic variations in the delivery of postacute rehabilitation services have been widely described in the USA.[3–6] For instance, these variations can occur across US Census Divisions[7, 8] and across rural versus urban areas[6, 9]. Geographic delivery variations often reflect variable supply of postacute care services or the need of people needing to travel long distances to access services[3, 9–14]. For example, IRFs and HHAs have been spatially segregated from rural hospitals, which might affect the delivery of postacute care services for patients with a rural residency[3]. Not surprisingly, patients residing in rural areas were less likely to be discharged to HHA care or IRFs than to SNFs[13, 14]. When patients were discharged to HHA care, rural residents received fewer rehabilitation therapy visits, and less so for those living alone and with comorbidities[15]. All the above compounds to higher rates of hospital readmissions and of adjusted 30-day mortality among rural versus urban residents[9, 15].

Among rural counties, those that have lower population size (fewer than 5,000 population) more often experience lower rehabilitation-therapy delivery rates.[8] However, an ecological fallacy can still apply[16, 17]. This means that some rural counties may not have low delivery despite having the ecological risk (e.g., lower population), and vice-versa. In turn, geographic variation across US Census Division variations also has been widely observed[7, 8]. Altogether, the use of Geographic Information Systems (GIS) can help map exactly which counties have had the lowest delivery of postacute rehabilitation therapies[18, 19]. GIS has been increasingly used in health research[20–22] to analyze the supply or delivery of health, rehabilitation or other support services for rural residents in Australia, Canada, Japan, or Brazil[18, 19, 23-27]. For the USA, GIS-based methods have been applied to one State, using data from 2013-2014[28].

To our knowledge, a specific form of GIS-based analysis, *hot* and *cold spot* analyses[22, 29, 30], has not been used to identify the geographic areas (e.g., clusters of counties that are geographically closer to each other) that have significantly higher or lower rates of rehabilitation therapy delivery compared to the remaining counties in the United States (US). Here, we aim to use *hot* and *cold spot* analyses to map which counties and clusters of adjacent counties are utilizing rehabilitation therapy services at the lowest rates. We used public-domain Original Medicare data to 1) map the risk-adjusted postacute therapy delivery rates by county; 2) identify and map the *cold spots* of those delivery rates; and 3) analyze whether those *cold spots* are more frequent in rural and small counties.

## Materials and methods

### Design

Statistical spatial clustering analysis through ArcGIS Pro (version 3.4, ESRI, Inc.,) software, using a *hot spot and cold spot* analysis, focused on identifying *cold spots* of rehabilitation therapy delivery. Cold-spots, in this case, represent clusters of areas with lower risk-adjusted delivery rates when compared to other regions of the US.

### Key Measures

We developed and used several measures. First, we computed therapy delivery volume, defined as the total number of therapy minutes provided to Original Medicare beneficiaries by type of postacute care provider located within each county. Then, using those county-level therapy delivery volume data, we computed therapy delivery rates, defined as therapy minutes per Original Medicare beneficiary in the county and adjusted those data using the Hierarchical Condition Category (HCC) risk score to account for differences in potential need for therapy.

Spatial clustering analysis then identified *hot spots* (clusters of statistically high delivery rates) and *cold spots* (clusters of statistically low delivery rates). We calculated the proportion of cold spots in rural and small rural counties, and their absolute and relative percentage-point differences from the national baseline (i.e., the proportion of U.S. counties that are rural or small rural). Additional details on how these measures were computed are provided in the subsections below.

### Computing county-level postacute therapy delivery volume

The Medicare Post-Acute Care and Hospice Provider Utilization and Payment Public Use File (PAC PUF)[31] was the main data source for computing the postacute therapy delivery volume, in minutes of therapy provision, for each US county. The PAC PUF contains provider-aggregated OriginalMedicare data for selected Part A covered postacute care facilities. Specifically, it provides the total documented minutes of physical therapy, occupational therapy and speech and language pathology, inclusive of assistants (e.g., physical therapy assistants), for each Medicare-certified SNF, HHA, and IRFs. The PAC PUF is updated annually, adding one additional year of data. Here, we used the version released in 2024, whose most recent year of data is 2022[31].

Although the PAC PUF contains data from Puerto Rico as a territory as well as from Alaska and Hawaii as states, here we only extracted and computed therapy delivery data from the 48 contiguous States and the District of Columbia because the geographic distances of the excluded locations to the other states would affect the statistical spatial clustering.

### Computing county-level postacute therapy delivery rates

To compute the county-level delivery rates, we summed the therapy minutes from each PAC provider type (i.e., SNF, HHA, IRF) for each US county. Then, we computed county-level delivery rates for each provider type based on the provider location. This was done by dividing the respective county-level therapy minutes per the number of Original Medicare beneficiaries of the respective counties for 2022. The latter was obtained from the Medicare Geographic Variation Public Use File (GV PUF)[32]. To account for varying county-level risks of therapy delivery, we risk-adjusted that delivery rate for the 2022 county-level Hierarchical Condition Category (HCC) risk score, also present in the GV PUF[32]. The HCC risk score has been used, along with other data, for determining capitated payments for Medicare Advantage beneficiaries and accounts for the expected annual healthcare utilization[7, 32].

### Missing or incomplete data in the underlying dataset

The minutes of therapy delivery data in the PAC PUF file are not exhaustive. For instance, for privacy protection, the file suppresses the therapy minutes for SNFs, HHAs, and IRFs that served 10 or fewer Original Medicare beneficiaries within a given year. This may occur for example for providers who were closing or opening operations within a given year. Providers with suppressed therapy minutes were a minority, and present across the rural-urban spectrum. For example, in 2022 only 0.05% of the PAC providers had values suppressed for the three therapy types, with 68% of them local in urban counties[8]. Also, the number of therapy minutes registered in the PAC PUF reflects prevalent CMS documentation requirements, which may not be exhaustive of all those therapy minutes actually delivered. The **supplementary appendix 1** provides details on how the therapy delivery data in the PAC PUF is obtained, including applicable limitations per provider type. For example, the reported therapy minutes for IRFs are the ones delivered during the first 14 days only; this is the average IRF length-of-stay[31]. Thus, therapy minutes provided in the 15^th^ or subsequent days for patients who had stayed longer than 14 days were not accounted for. For SNFs, therapy minutes reflect only the claims paid under the SNF Prospective Payment System, which excludes any “swing beds” in Critical Access Hospitals. For SNFs, the therapy minutes are not exhaustive (e.g., not included when the patient died during the stay). Nonetheless, the same documentation requirements applied among providers of the same type, regardless of geography, enabling the relative delivery-rate comparisons across geographies.

### Visualization: choropleth maps

To visualize the computed delivery rates, we used ArcGIS Pro to map the county-level delivery rates as *choropleth* maps. These maps display the county-level delivery rates (HCC-adjusted), classified into five gradients in addition to true zeros (i.e., a sixth gradient for counties with no postacute therapy minutes documented in the PAC PUF). The five gradient bins were determined at 20 percentile-rank intervals of the HCC-adjusted delivery rates, so that each gradient contains the same number of counties.

### *Hot* & *cold spot* analysis and rural-urban comparison

The *hot spot analysis* tool in ArcGIS Pro was used to identify and map clusters of high (hot spots) and low (cold spots) postacute therapy delivery rates. *Cold spots* refer to statistically significant clusters of low values, while *hot spots* are clusters of high values. This analysis is based on a computed *Getis-Ord Gi\** statistic for each county and compares local averages with national averages to identify statistically significant clusters of high and low values. The output generated from the *G_i_*^∗^ statistic consists of standardized *z*-scores and can be used to determine the statistical significance of the *G_i_*^∗^ value. A positive *Gi\** statistic with a significant *p* value indicates intensity of clustering around high values (i.e., *hot spot*), while a negative *Gi\** statistic with a significant *p* value indicates clustering of low values (i.e., *cold spot*). Gi* values approaching zero and with non-significant *p* values imply no spatial clustering, or random distribution of the observed rates (i.e., neither *hot* nor *cold spots* of delivery rates). Spatial relationships among counties were defined using a fixed-distance spatial conceptualization. This approach was selected to ensure consistent spatial interaction across counties of highly variable geographic size and population density. All counties within the specified distance band were treated as neighbors and equally weighted in the analysis. No classification methods (e.g., natural breaks) were used for the clustering step, as *hot* and *cold spots* were identified based on the statistical significance of *Gi* z-scores*.

To reduce the impact of outlier rates in the *hot* and *cold spots* determination, particularly in small-size rural populations with low denominators,[8] we log transformed the HCC-adjusted delivery rates prior to analysis. The *hot* and *cold spots* were identified and mapped with 90%, 95% and 99% confidence intervals to illustrate the full range of statistical significance levels within a single overlay.

We initially conducted a “general”[33, 34] *hot spot* analysis, in which counties were used as our spatial administrative boundaries. However, for robustness, we also ran an “optimized” *hot spot* analysis,[35-37] in which no conception of spatial relationships is defined prior to running the analysis. In the latter way, the software interrogates the data to determine which geographies and their aggregation optimize the analytical results, determining the appropriate scale and correcting for spatial dependence and multiple testing[35, 36].

Finally, the number and percentage among all counties of statistically significant *cold spots* identified by the “general” *hot spot* analysis (at 95% confidence level) were mapped and compared against two rural classification systems and the percentage rural counties according to the given classification. The first classification system was a binary rural-urban measure derived from the 2023 Core-Based Statistical Areas (CBSA)[31], which is used by the Centers of Medicare & Medicaid Services to define rural residency of their beneficiaries and for overall payment policies[31]. The second classification focused on identifying “small rural” counties, defined as those with the smallest population size (Rural-Urban Continuum Codes 8 and 9). This category represents rural contexts found to have the highest ecological risk of having lower risk-adjusted postacute delivery rates[8].

## Results

### Choropleth maps

National (excluding Alaska and Hawaii), the county-level mean (SD) HCC-adjusted therapy delivery rates for HHAs, IRFs, and SNFs were 30.3 (±58.8), 7.7 (±26.2), and 66.5 (±52.9) minutes per beneficiary, respectively. **Figures 1 to 3** provide *choropleth* maps visually depicting the county-level HCC-adjusted delivery rates into gradient levels, respectively for SNFs, HHAs, and IRFs. In Figure 3 for IRFs, we observe many counties without a colored gradient, which means the HCC-adjusted delivery rates were equal to zero, likely due to the absence of IRFs in the given counties. In **Figures 1 and 2**, those uncolored gradients were less prevalent overall, but still often concentrated in States with larger rural populations, such as Montana, North and South Dakota.

**Figure 1:**
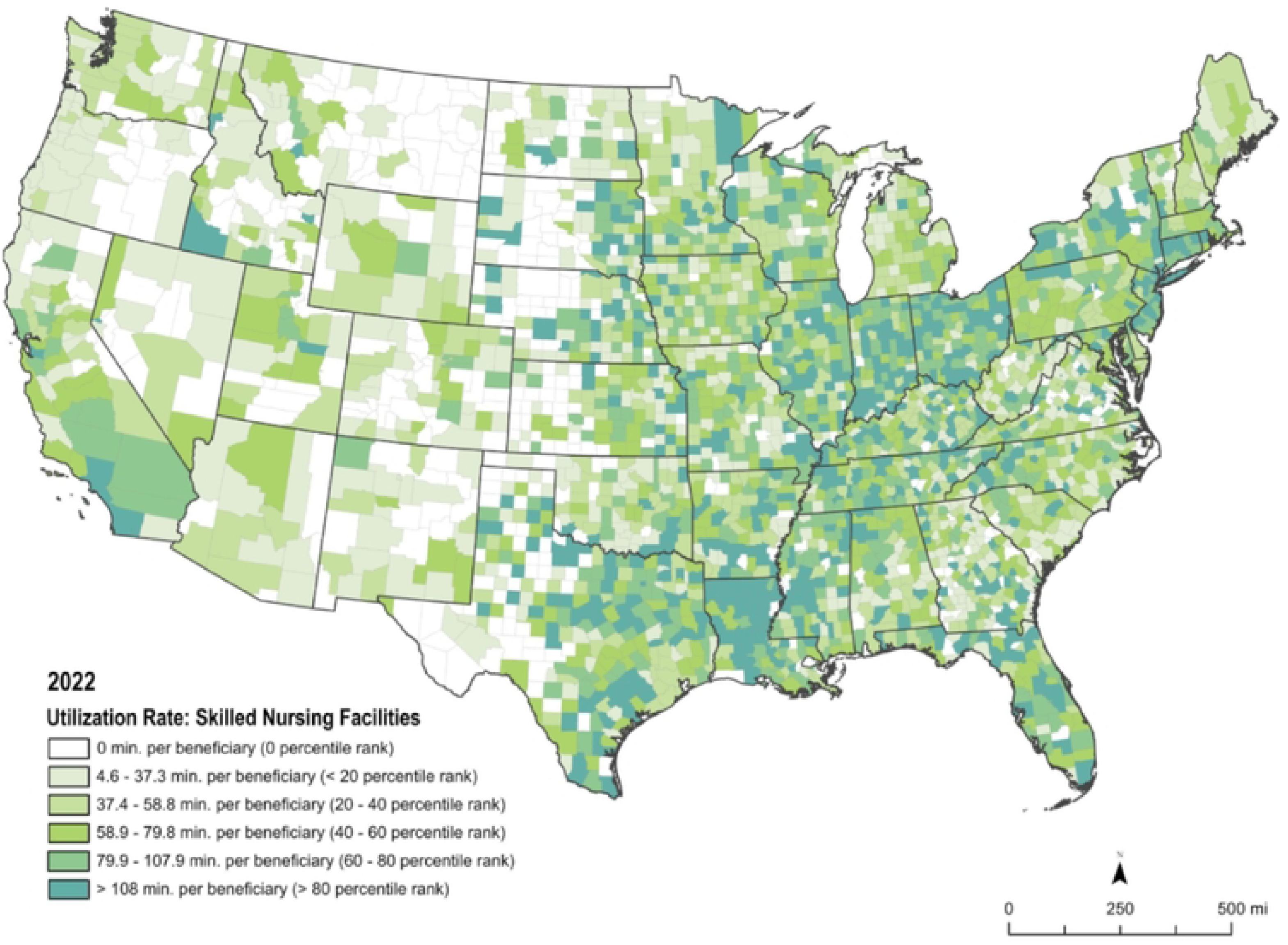
Choropleth map of the therapy delivery rate for Skilled Nursing Facilities (SNFs), adjusted for the Hierarchical Condition Category risk score. County and state spatial boundaries were sourced from the U.S. Census Bureau [2020].

**Figure 2:**
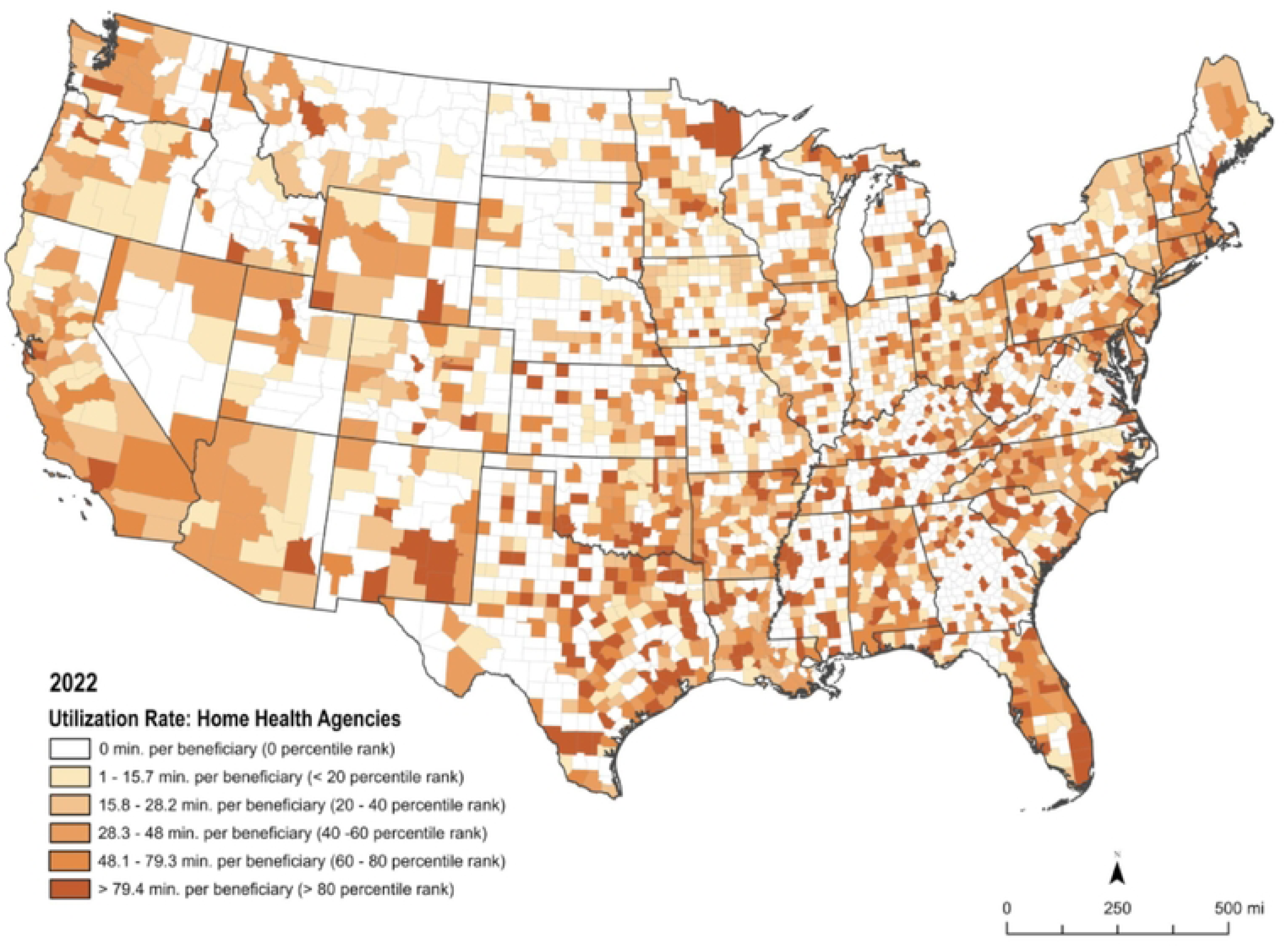
Choropleth map of the therapy delivery rate for Home Health Agencies (HHAs), adjusted for the Hierarchical Condition Category risk score. County and state spatial boundaries were sourced from the U.S. Census Bureau [2020].

**Figure 3:**
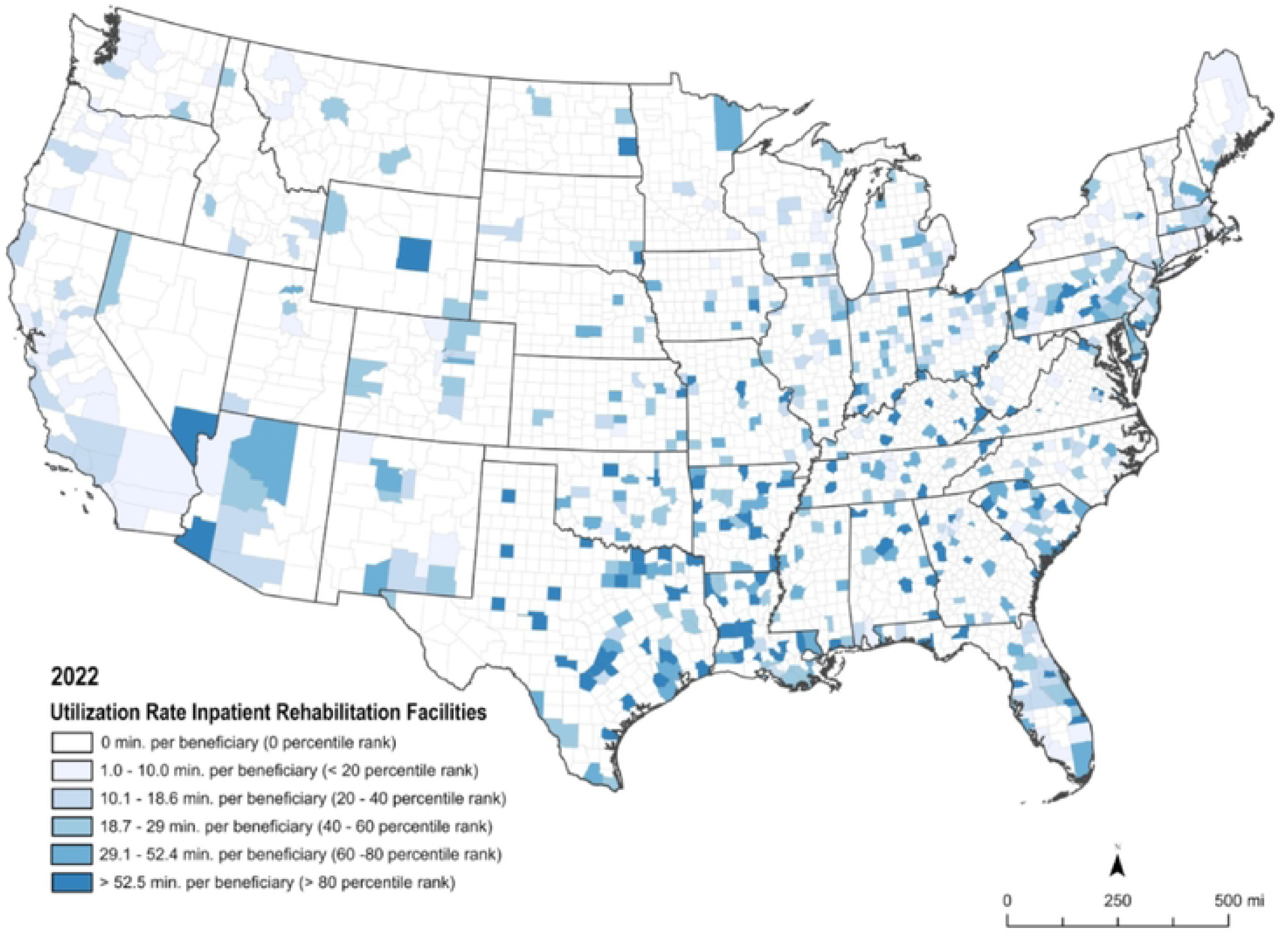
Choropleth map of the therapy delivery rate for Inpatient Rehabilitation Facilities (IRFs), adjusted for the Hierarchical Condition Category risk score. County and state spatial boundaries were sourced from the U.S. Census Bureau [2020].

### Statistical spatial clustering – *cold spot* analysis

Across the 3,108 included counties (excluding Alaska and Hawaii), *cold spots* of therapy delivery rates at the 95% confidence level were identified in 17%, 16%, and 9% of counties for SNFs, HHAs, and IRFs, respectively, based on the “general” hot and cold spot analysis. In turn, *hot spots* were identified in 36%, 15%, and 12% of counties respectively for the same provider types and analysis. The remaining counties were not identified as statistically significant *hot* or *cold spots* at the 95% confidence level.

**Figures 4, 5 and 6** provide map results for the “general” *hot* and *cold spot* analysis. In turn, **Supplementary Figures 1, 2, and 3** provide the “optimized” version (i.e. with no administrative boundaries inputted) of *hot* and *cold spot* analysis respectively for SNFs, HHAs, and IRFs.

In Figure 4, for SNFs, we observed large *cold spots* particularly in the western side of the West North Central and West South-Central US Census Divisions, and within the Mountain Division. The latter extended towards the eastern part of the Pacific division.

In Figure 5, for HHAs, the cold spots were spread across various US Census Divisions. There were areas in the East North Central and East South Central (e.g., parts of Kentucky, Indiana or southern Illinois) that were *cold spots* for HHAs which were *hot spots* for SNFs. Many counties in Georgia were also cold spots for HHAs.

Finally, in Figure 6 for IRFs, *cold spots* were especially prevalent in the West North Central, with some pockets elsewhere (e.g. Geogia, Mississippi, Kentucky, West Virgina), in the context of many counties without reported minutes for IRFs (figure 3).

**Figure 4:**
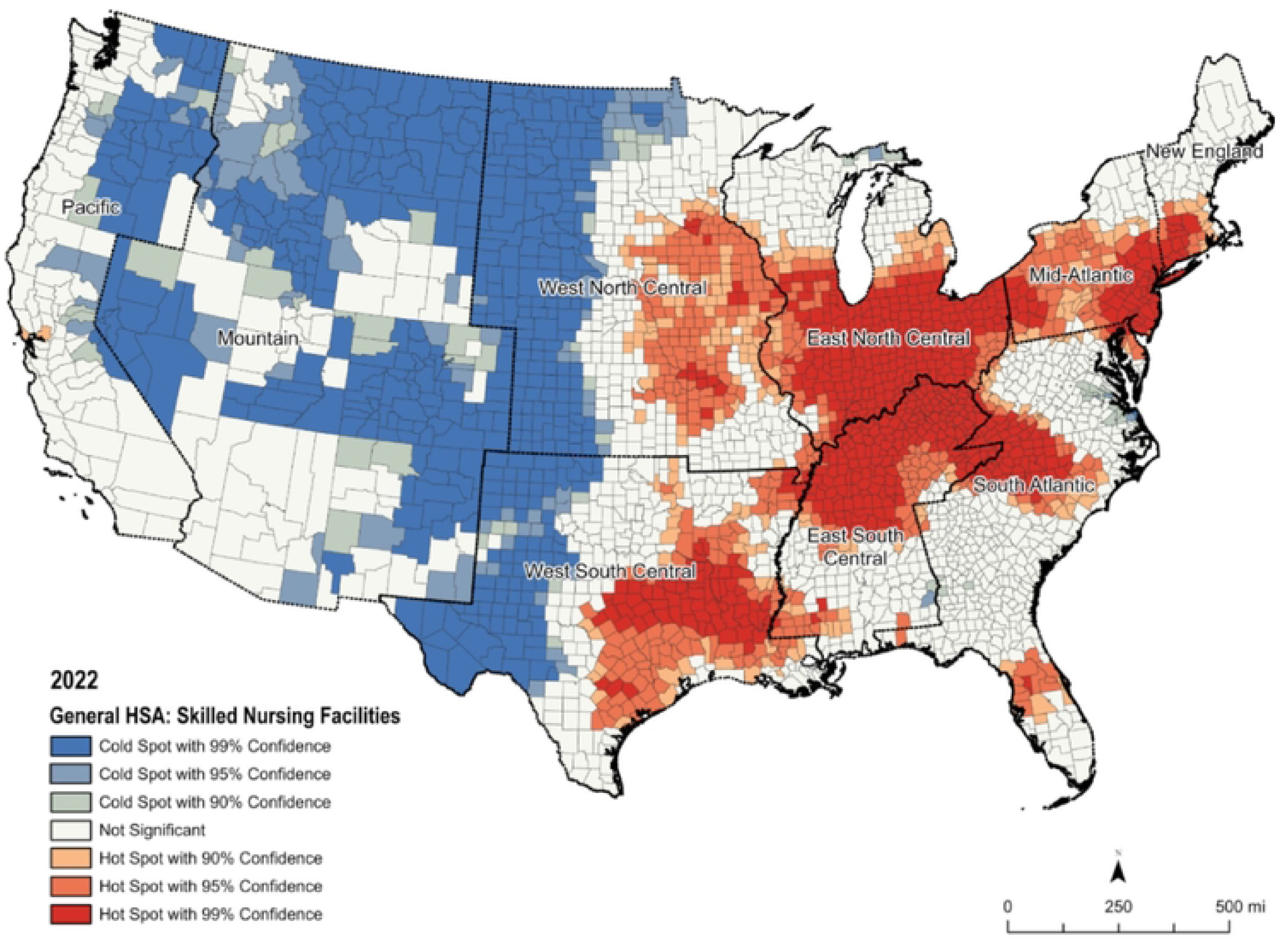
General *hot spot* and *cold spot* analysis of the therapy delivery rate for Skilled Nursing Facilities (SNFs),, adjusted for the Hierarchical Condition Category risk score, with bolder demarcation and label (green) for the nine US Divisions. County and US Division spatial boundaries were sourced from the US Census Bureau [2020].

**Figure 5:**
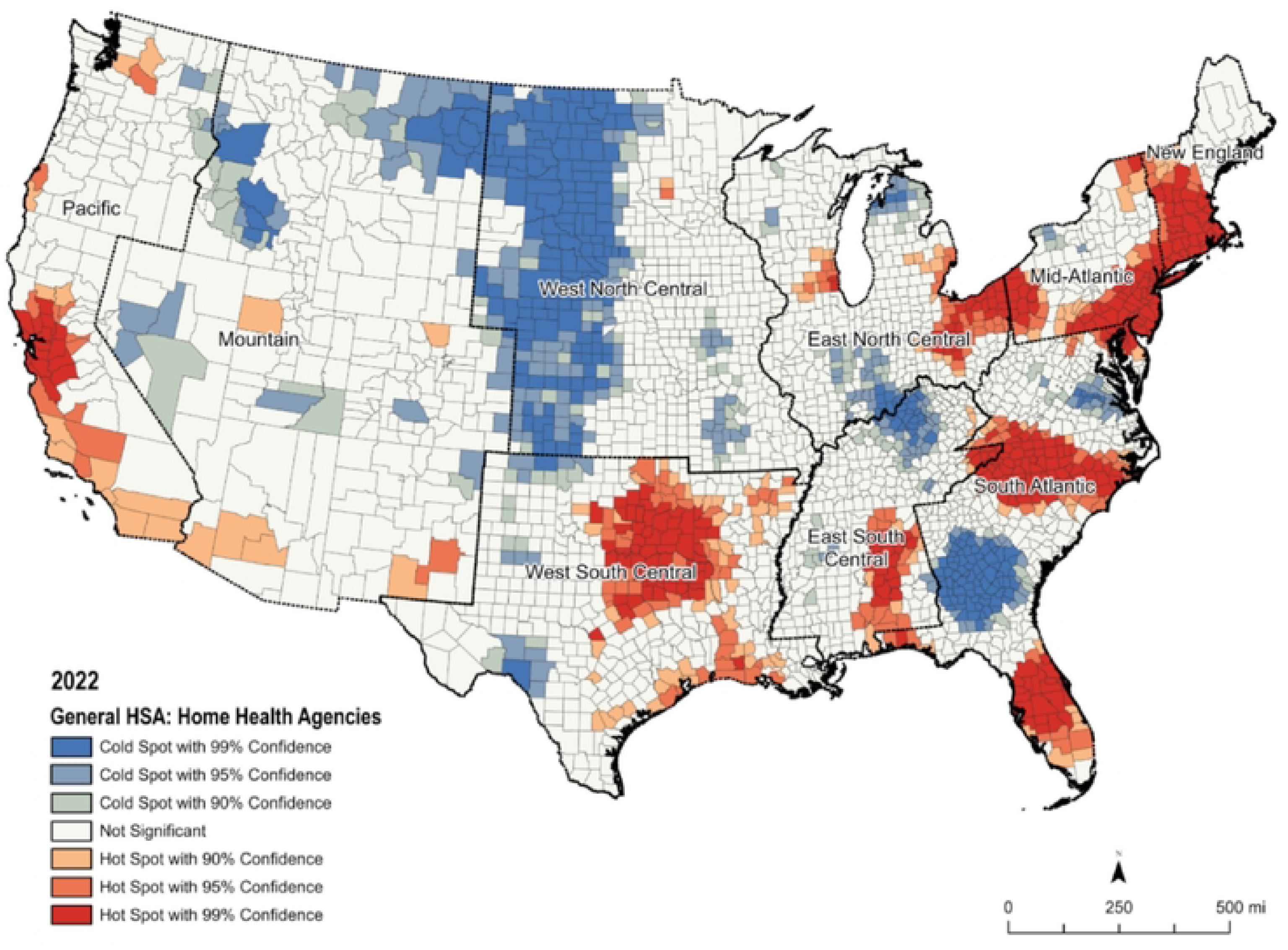
General *hot spot* and *cold spot* analysis of the therapy delivery rate for Home Health Agencies (HHAs),, adjusted for the Hierarchical Condition Category risk score, with bolder demarcation and label (green) for the nine US Divisions. County and US Division spatial boundaries were sourced from the US Census Bureau [2020].

**Figure 6:**
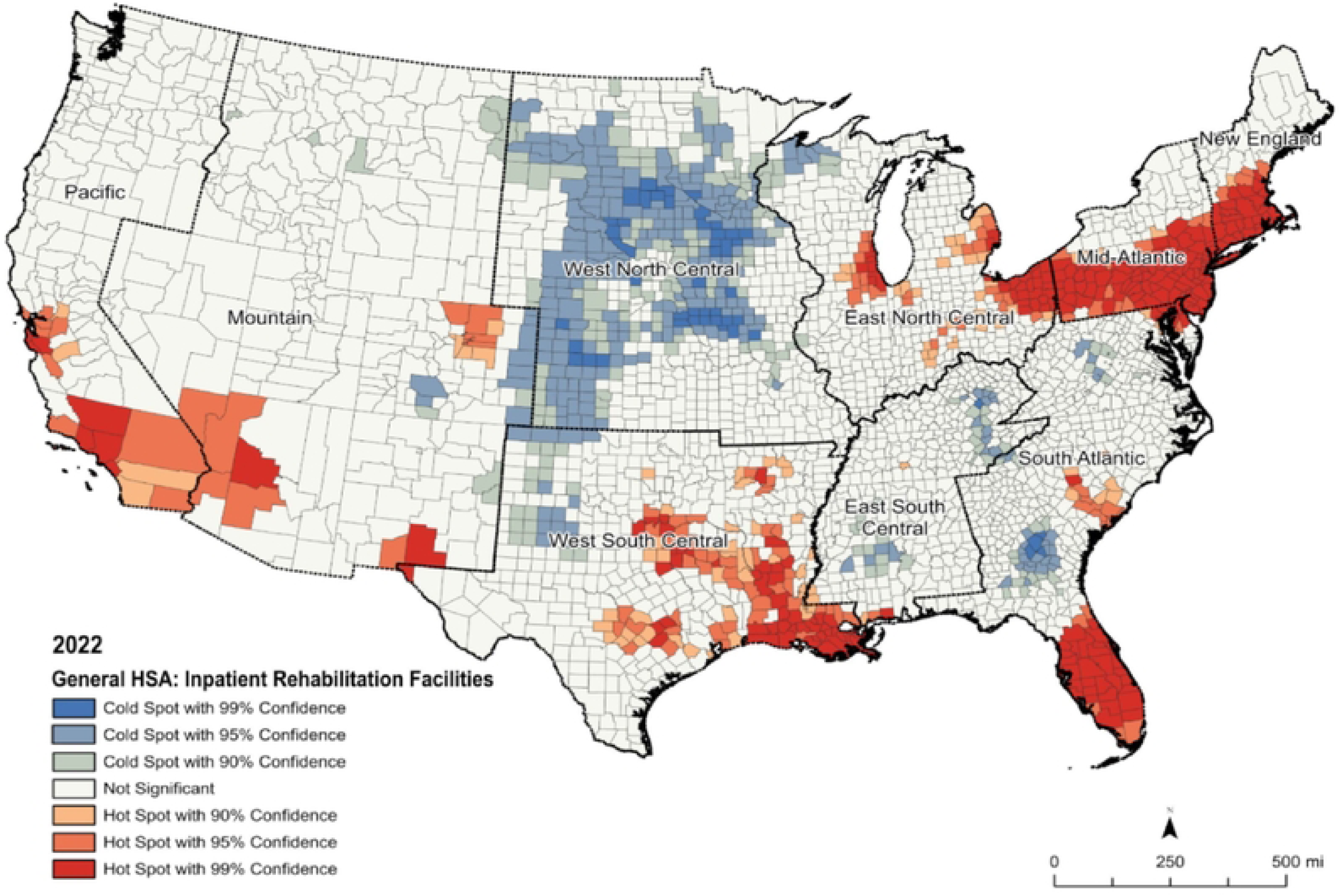
General *hot spot* and *cold spot* analysis of the therapy delivery rate for Inpatient Rehabilitation Facilities (IRFs), adjusted for the Hierarchical Condition Category risk score, with bolder demarcation and label (green) for the nine US Divisions. County and US Division spatial boundaries were sourced from the US Census Bureau [2020].

Finally, the “optimized” versions of the analysis (**supplementary figures 1, 2, and 3**) identified the large landmasses of *hot* and *cold spots* also overall identified above, but less so for relatively confined pockets of *cold spots* identified in the “general” *hot* and *cold spot* analysis.

### *Cold spot* prevalence in rural areas

**Figure 7** shows that the percentage of *cold spots* located in “rural” counties is 58.7% for HHAs, 63.6% for SNFs, and 70.5% in IRFs, whereas only 41.1% of U.S. counties (excluding Alaska and Hawaii) are classified as rural. Thus, the percentage of *cold spots* in rural counties exceeds the percentage of rural counties by 17.6, 22.5, and 29.4 percentage points, respectively for HHAs, SNFs and IRFs. When expressed as percent change (i.e., percentage point difference divided by the baseline), the percentage of *cold spots* in rural counties is 42.8%, 54.7% and 71.5% higher than the percentage of “rural” counties in the USA. This percent change metric enables a parallel comparison with the “small rural” counties reported in the following paragraph, which have a lower baseline.

**Figure 8**, in turn, focused on “small rural” counties and shows similar trends, though with even larger differences. The percentage of *cold spots* located in “small rural” counties is 56.3% for HHAs, 59.4% for SNFs, and 64.1% for IRFs, while only 33.3% of counties in the USA are classified as “small rural” under this system. Thus, the percentage of cold spots in “small rural” counties exceeds the percentage of “small rural” counties by 23.0, 26.1, and 30.9 percentage points, respectively, for HHAs, SNFs, and IRFs. Expressed as percent change, the percentage of cold spots in small rural counties is 69.1%, 78.4%, and 95.5% higher than the percentage of counties classified as small rural in the USA.

**Figure 7:**
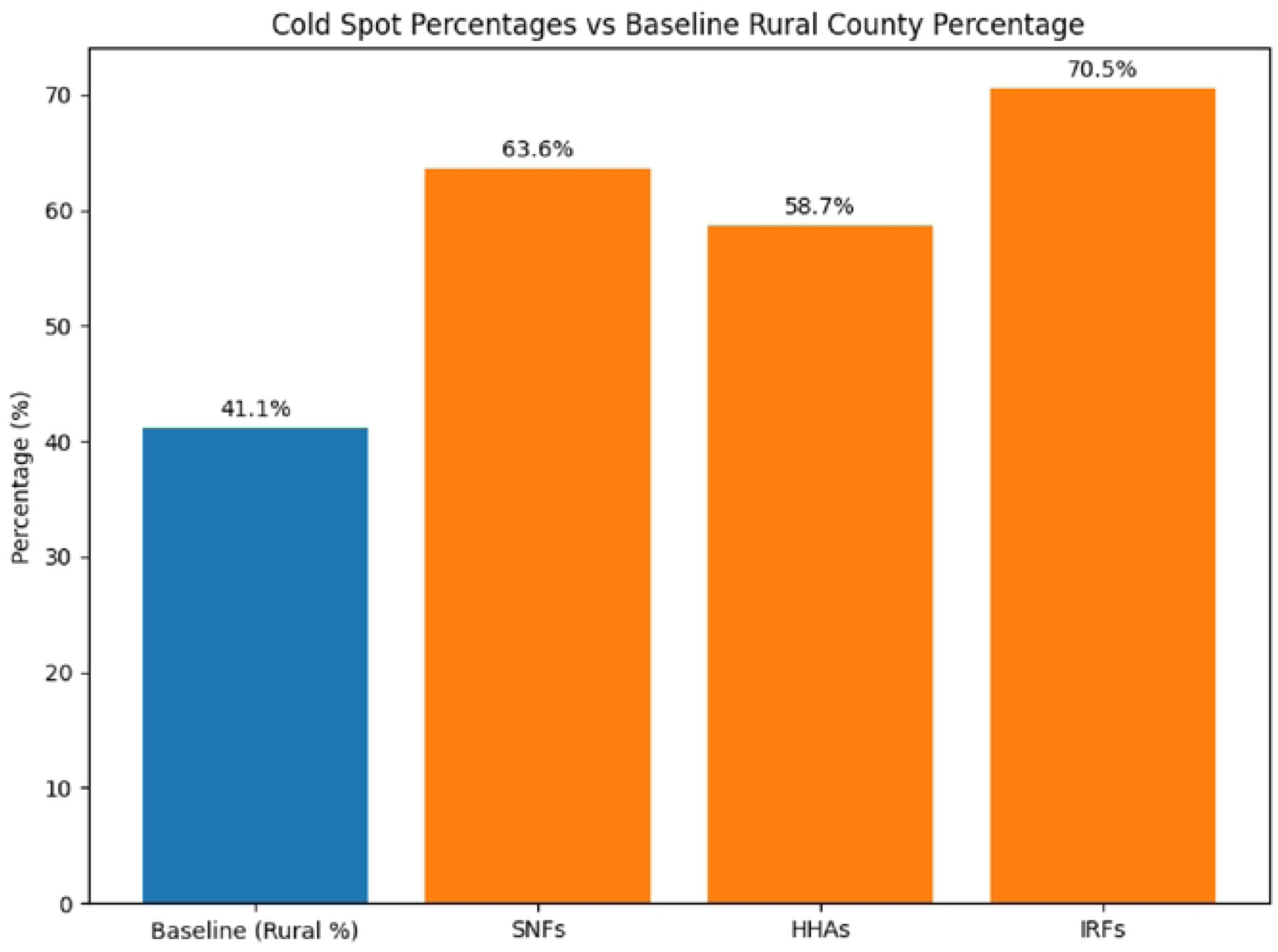
Percentage of cold spots located in *rural* counties (CBSA classification) – in bolded black numbers, including the percentage points above and beyond the percentage of *rural* counties in the USA according to the same rural county classification. Data for SNFs, HHAs, and IRFs. Notes: The *cold spots* used for computing these percentages were those identified by the general *hot* and *cold spot* analysis with a 95% Confidence Interval (CI). The USA counties here exclude those from Alaska and Hawaii Legend: SNF: Skilled Nursing Facilities; IRF: Inpatient Rehabilitation Facilities; HHAs: Home Health Agencies. CBSA: Core-based Statistical Areas.

**Figure 8:**
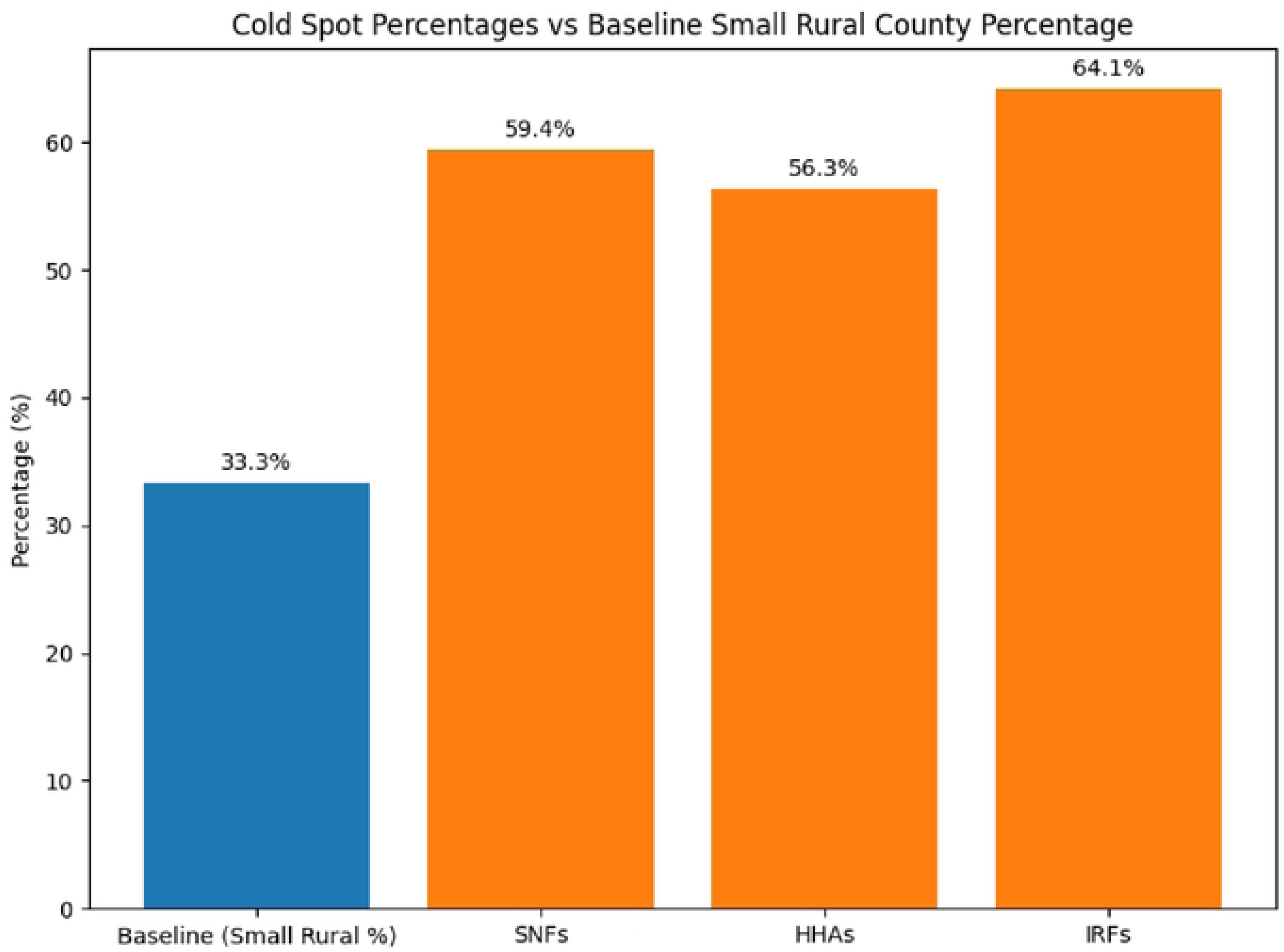
Percentage of cold spots located in *small rural* counties (grades 8 and 9 of the RUCC classification) – in bolded black numbers, including the percentage points above and beyond the percentage of rural counties in the USA according to the same *small rural* county classification. Data for SNFs, HHAs, and IRFs. Notes: The *cold spots* used for computing these percentages were those identified by the general *hot* and *cold spot* analysis with a 95% Confidence Interval (CI). The USA counties here exclude those from Alaska and Hawaii Legend: SNF: Skilled Nursing Facilities; IRF: Inpatient Rehabilitation Facilities; HHAs: Home Health Agencies. RUCC: Rural-Urban Continuum Codes.

## Discussion

This paper used GIS-based methods to map and spatially analyze which counties, and clusters of counties, utilized rehabilitation therapy services the least, based on public-domain, provider-aggregated Original Medicare data for 2022. We mapped *cold spots* of HCC-adjusted rates of documented therapy minutes for SNFs, HHAs, and IRFs. *Cold spots* varied by setting type. For example, SNFs *cold spots* were especially identified in the Mountain and West North Central US Census Division, while HHAs *cold spots* were distributed across more US Divisions.

Importantly, *cold spots* of postacute therapy delivery rates were more prevalent in rural counties and especially in rural counties with smaller population size, across site types.

The mapped delivery trends of HHAs and SNFs displayed some similarities, as well as some unique or even opposing patterns. For example, some parts of Kentucky, Indiana or southern Illinois were *cold spots* for HHAs, while *hot spots* for SNFs. The lower availability and delivery of HHAs may lead to greater use of rehabilitation in SNFs - where these are available and the former might not. That may have negative consequences. For instance, the lack of local supply for HHAs may affect the residents’ opportunities for “ageing in place”, which is preferable from health, social and economic points of view[38]. The recently ceased rural-add on for rural HHAs removes a compensatory incentive for HHAs to operate in rural counties, while it was effective when in place and of enough size (5%-10%)[9]. Hence, the greater likelihood of *cold spots* of HHA therapy delivery in rural and small rural areas observed in 2022 may prevail or be aggravated in the years to come without target policy changes.

For IRFs, the choropleth maps showed many counties and large geographies with no reported minutes. IRFs provide more intensive therapy which often leads to better functional outcomes for people recovering from a stroke[1]. But IRFs often cluster around medical academic centers or areas with larger population density[3]. Populations with no geographical access to IRFs need to rely on SNFs which often provide less intensive therapy, even if the patient can benefit from greater therapy intensity. Hence, SNF delivery in rural and other counties absent from an IRF may include service to patients that otherwise would be served by IRFs[4, 28]; nonetheless, *rural* and *small rural* counties were still more likely cold spots of delivery for SNFs.

Many rural and especially small rural and remote locations refer to low-density markets with low economic attractiveness to operate with fixed costs[39-41]. Hence, alternative or supplementary models of rehabilitation service delivery may be needed. For example, telerehabilitation options showed overall cost-effectiveness and satisfaction[42, 43], but its uptake has been suboptimal and highly variable due to personal and contextual factors such as attitudes toward telehealth of both the therapists and clients[44-46], as well as the lower internet bandwidth in rural and remote areas[44, 45, 47-49]. Global efforts have increasingly emphasized the integration of rehabilitation services within primary care, particularly in underserved settings, to address the needs of ageing populations through more proximal, community-based care model[50, 51].

Similar approaches could be considered within the U.S., particularly in rural and underserved areas. Furthermore, rural practitioners working in large catchment areas or those addressing a wide range of health conditions often need to be remotely supported by provider-to-provider continuous education and consultation from more specialized health professionals in a hub (e.g., Project ECHO® model[52, 53], also being applied to rehabilitation[54, 55]). These or other forms of providing or supplementing rehabilitation delivered at where people live might be considered, especially for large geographic areas with no or minimal providers or “zero” delivery rates.

Finally, it is noteworthy that our *hot* and *cold spots* had a US national focus and were therefore defined relative to nationwide values; accordingly, different results may be obtained if analyses are conducted at the regional, division, or state level instead of nationally. For instance, the presence of many counties with low or zero therapy delivery in the Mountain and West North

Central U.S. divisions lowers the national aggregate and widens the overall dispersion, which can affect the analytical sensitivity to detect cold spots in other regions. Accordingly, future GIS-based research could benefit from analyses conducted at regional or state levels to identify within-region and within-state variation, complementing the nationwide comparisons presented here and more directly informing state and local policy and programmatic decisions. In addition, because state-level clustering analyses may be constrained by the relatively small number of counties in many states—potentially limiting the stability of spatial clustering results—future studies may benefit from the use of more granular geographic units (e.g., ZIP Code Tabulation Areas or Census tracts) to support robust sub-state or regional analyses.

### Limitations

This study has several limitations. First, the mapping and geospatial analysis reflect the limitations of the underlying dataset. The therapy minutes reported were not exhaustive of each patient’s entire length-of-stay, and suppressed values existed for providers serving 10 or less Original Medicare in 2022. These limitations apply across geographies, enabling here the relative geographic analyses and comparisons. Second, the delivery rates we used from the public-domain dataset reflect the county where the provider is based, not necessarily where the patient resides. Further analysis should account for the ‘traveler’ effect (e.g., patients that cross county borders to obtain postacute care). Third, the geospatial analysis could not be applied to all states, for example, Hawaii and Alaska, due to geographic constraints. Fourth, the dataset refers only to beneficiaries with Original Medicare for postacute care Part A services, not Medicare Advantage beneficiaries, commercially insured, or Medicaid-only populations. Fifth, the data do not account for rehabilitation provided in long-term care hospitals or therapy in rehabilitation units in

Maryland not classified as IRFs, therapy provided under Medicare Part B including long-term residents of nursing homes and outpatient rehabilitation. Sixth, the PAC PUF dataset does not include therapy delivery data from swing beds in Critical Access Hospitals; therefore, the rural SNF delivery estimates presented here reflect care delivered through the more stable SNF service capacity in rural areas. Seventh, there is no standard on the ‘right-size’ therapy delivery rate, which means that low or high delivery results do not necessarily equate to an under- or over-delivery, rather greater or lower delivery rates compared to the overall national trend. Finally, we did not present a combined measure or analyses across postacute care provider types (i.e., county-level delivery rates combining HHAs, SNFs, and IRFs) because differences in data completeness across provider types in the underlying dataset, together with the uneven geographic distribution of provider types, would introduce measurement bias at the county level, compromising comparability across counties (including rural–urban comparisons) and the validity of spatial clustering results.

## Conclusion

*Cold spots* of postacute rehabilitation therapy delivery varied across the continental USA per provider type but were especially prevalent in rural and small rural counties ─ across all provider types. Knowing where these *cold spots* are located may inform geographically-target policies. That may include alternative service delivery options to supplement postacute rehabilitation services where these are not available or economically viable due to very low-density markets.

## Data Availability

All data used in this study are derived from publicly available from the Centers for Medicare & Medicaid Services (CMS), including the Medicare Post Acute Care and Hospice Provider Utilization and Payment Public Use File (PAC PUF) and the Medicare Geographic Variation Public Use File (GV PUF). Our aggregated data is also openly available in the Openicpsr at: https://www.openicpsr.org/openicpsr/project/233575/version/V2/view

## Acknowledgments

None.

## Captions (Supplementary Figures)

**Supplementary Figure 1**: Optimized *hot spot* and *cold spot* analysis of the therapy delivery rate in 2022 for Skilled Nursing Facilities (SNFs), adjusted for the Hierarchical Condition Category risk score. County and state spatial boundaries were sourced from the US Census Bureau [2020].

**Supplementary Figure 2**: Optimized *hot spot* and *cold spot* analysis of the therapy delivery rate in 2022 for Home Health Agencies (HHAs), adjusted for the Hierarchical Condition Category risk score. County and state spatial boundaries were sourced from the US Census Bureau [2020].

**Supplementary Figure 3**: Optimized *hot spot* and *cold spot* analysis of the therapy delivery rate in 2022 for Inpatient Rehabilitation Facilities (IRFs), adjusted for the Hierarchical Condition Category risk score. County and state spatial boundaries were sourced from the US Census Bureau [2020].

